# Mental health and health behaviours in adolescence and risk of being NEET from ages 16 to 24: longitudinal findings from the UKHLS

**DOI:** 10.64898/2026.08.12.26360290

**Authors:** Zixu Li, Jacques Wels, Nishi Chaturvedi, Praveetha Patalay

## Abstract

**Background:** Young people who are Not in Education, Employment, or Training (NEET) represent a major public health and societal challenge. Existing evidence has linked adolescent mental health problems and health risk behaviours to NEET but has largely treated NEET as a static, rather than longitudinal outcome and overlooked the combined effects of multiple health conditions.

**Methods:** Using data from 5,262 participants born between 1993 and 2000 in the UK Household Longitudinal Study, this study examined the independent and combined associations of adolescent mental health problems (emotional symptoms, conduct problems, hyperactivity) and health risk behaviours (regular smoking, drug use, alcohol use, and high social media use) with ever-NEET status, NEET chronicity, and NEET trajectories from ages 16 to 24, using modified Poisson, proportional odds, and multilevel logistic regression models, respectively.

**Findings:** All mental health problems were associated with ever-NEET status (RRs 1.24-1.27) and NEET chronicity (ORs 1.41-1.98); emotional symptoms showed a widening disadvantage with age, while the disadvantages associated with conduct problems and hyperactivity remained stable. Among health risk behaviours, regular smoking showed the strongest and most persistent relationships with NEET (ever-NEET RR 1.54; chronicity OR 1.64); drug use was related to ever-NEET status (RR 1.37) and an increasing disadvantage after age 21-22, while alcohol use and social media use showed limited associations. NEET risk generally increased with the number of co-occurring conditions, but for recurrent NEET (three or more occasions), risk was only elevated at three or more co-occurring conditions.

**Interpretation:** Adolescent health exposures were associated with NEET risk during ages 16-24, but the strength and pattern varied by exposure and outcome, offering potential insights into the timing and emphasis of any interventions.

## INTRODUCTION

Young people who are Not in Education, Employment, or Training (NEET) experience substantial social, economic, and health disadvantages, with broader societal consequences including reduced economic productivity, widening social inequalities, and increased demands on public resources.^1^ Despite the Sustainable Development Goal (SDG) target to reduce NEET by 2020, global rates have remained persistently high at around one in five young people over the past decade.^2^ In the UK, NEET rates have risen steadily in recent years, reaching an estimated 13.5% among young people aged 16-24 years in early 2026, equivalent to more than one million young people.^3^

Meta-analytic evidence suggests that mental health problems (e.g., mood disorders and behavioural problems) and health risk behaviours (e.g., drug and alcohol use) are associated with an elevated risk of NEET status; however, findings become less consistent when disaggregated, with subgroup analyses further suggesting that these associations vary across age groups and conditions.^4,5^ This heterogeneity likely reflects the dynamic nature of the transition to adulthood, during which both NEET risk and its associations with mental health problems and health risk behaviours are likely to vary. Yet, existing evidence has predominantly examined NEET outcomes at a single time-point or treated it as a static risk,^1,4^ thereby overlooking how these transitions unfold across adolescence and young adulthood. A life-course perspective on how risk factors in earlier life shape NEET trajectories is therefore needed to identify critical windows for prevention and timely intervention.

A further limitation is that most existing studies of mental health and NEET status in the UK have examined single mental health conditions.^1,6–8^ Differences in study samples and methodologies make it difficult to compare the relative contribution of different mental health conditions to NEET risk within a common analytical framework, limiting the identification of young people at greatest risk and the delivery of targeted support. In addition, mental health problems and health risk behaviours frequently co-occur during adolescence;^9^ therefore, their independent and combined associations with NEET risk need to be examined, as co-occurring problems may confer additional risk.

Given these gaps, we adopted both life-course and cumulative-risk perspectives to examine the associations of adolescent mental health and health-risk behaviours with subsequent NEET outcomes. Specifically, using data from a UK household panel study, we longitudinally investigated whether mental health and health-related behaviours in mid-adolescence, individually and jointly, related to ever experiencing NEET status, NEET chronicity, and NEET trajectories between ages 16 and 24 years.

## METHODS

### Data source and analytic sample

The data used in this study were extracted from the UK Household Longitudinal Study (UKHLS), also known as Understanding Society.^10^ Understanding Society is a large-scale, nationally representative longitudinal survey of approximately 30,000 UK households that began in 2009 and has completed 15 waves to date (up to 2025). Respondents aged 16 and over complete the adult survey, whilst young people aged 10-15 complete a youth questionnaire.

The analytic sample was restricted to respondents who participated in both adult and youth surveys and had at least one valid NEET record between ages 16 and 24, with at least one valid exposure observation during adolescence. The sample was further limited to those born between 1993 and 2000, as respondents born after 2000 had not yet reached age 24 at the final wave of data collection. This resulted in a final analytic sample of 5,262 respondents and 25,539 person-observations of NEET status. Details of sample derivation are provided in Figure S1.

### Measures

#### NEET outcomes

NEET was derived from the adult survey labour force status variable. Respondents were classified as NEET if they were unemployed, long-term sick or disabled, or reported doing something else. All other labour force statuses were classified as non-NEET, including self-employment, paid employment, retirement, maternity leave, family care responsibilities, full-time education, participation in government training schemes, unpaid family work, apprenticeships, furlough, temporary layoff, and parental or adoption leave.

Based on repeated measures of NEET from all available waves between ages 16 and 24 years, we derived two additional outcome variables. First, ever NEET was defined as being observed in NEET status at least once during this age range. Second, among participants who experienced NEET, we calculated the cumulative number of NEET occasions and categorized it into three ordered groups: one, two, and three or more occasions.

***Adolescence exposures*** were extracted from the youth surveys completed by household members aged between 10 and 15 years. As some exposure variables were not measured at every wave, we assigned each respondent their most recent available observation. The age at which this observation occurred was highly concentrated across all exposures, with median values of 14-15 years (IQR 14-15).

Emotional symptoms, conduct problems, and hyperactivity were measured using corresponding subscales of the Strengths and Difficulties Questionnaire (SDQ), with higher scores indicating greater difficulties. Established cutoffs to identify clinical levels of symptoms were used: >=7 for emotional symptoms, >=5 for conduct problems, and >=7 for hyperactivity.^11^

Regular smoking was derived from questions on ever smoking and smoking frequency. Participants who had never smoked, had smoked only once or twice, or were former smokers were classified as non-regular smokers. Those who reported smoking sometimes, weekly, or more than six cigarettes per week were classified as regular smokers.

Drug use was derived from questions on glue/solvent sniffing, cannabis use, other illegal drug use, and frequency of illegal drug use. Participants were classified as drug users if they reported any use of glue or solvents, cannabis, or other illegal drugs, or reported using illegal drugs at least once. Those reporting no use across all drug-related measures were classified as non-users. Participants with incomplete or inconsistent responses were coded as missing.

Weekly alcohol use was derived from two questions: whether participants had ever consumed alcohol, and the frequency of alcohol consumption in the past four weeks, originally measured on a 5-point scale (1 = most days, 2 = once or twice a week, 3 = 2-3 times, 4 = once only, 5 = never). Participants who had never consumed alcohol or reported drinking less than weekly were classified as non-weekly drinkers, whereas those who reported drinking at least weekly were classified as weekly drinkers.

Heavy alcohol use was derived from questions on lifetime alcohol use and the number of intoxication occasions during the past four weeks (1 = none to 7 = 40 or more occasions).

Participants who had never consumed alcohol or reported no intoxication during the past four weeks were classified as non-heavy drinkers. Those who reported one or more occasions of intoxication were classified as heavy drinkers.

High social media use was derived from questions on computer access, social networking site membership, and weekday social media usage. Participants were classified as high users if they reported 4 or more hours of weekday social media use, consistent with previous research linking this level of use to poorer emotional health and behavioural difficulties in UK adolescents.^12^ All other participants, including those with no access to social media (no computer access at wave A or no social networking site membership) or 3 hours or less of weekday use, were classified as low users. Weekday social media use was originally assessed by asking: “On a normal weekday (Monday to Friday), how many hours do you spend chatting or interacting with friends through social media, gaming websites or apps?” Responses were recorded on a 5-point scale (1 = none to 5 = 7 or more hours per day).

Co-occurrence of mental health problems and health risk behaviours was operationalized using a cumulative risk score. The score was calculated by summing eight binary indicators: emotional symptoms, conduct problems, hyperactivity symptoms, regular smoking, drug use, weekly alcohol use, heavy alcohol use, and high social media use. Each indicator was coded as 1 if present and 0 otherwise. The resulting score ranged from 0 to 8 and was categorized into four groups: 0, 1, 2, and 3+ co-occurring conditions.

#### Covariates

Sex (male/female), birth year, and ethnicity (white/ethnic minority) were included as demographic covariates. Region of residence (London and the South of England, Midlands and North of England, Northern Ireland, Scotland and Wales), equivalised household income, and area deprivation (Index of Multiple Deprivation, IMD) during adolescence were also adjusted for, using each respondent’s most recent available observation from the youth survey to align with exposure measurement. Equivalised household income was derived from monthly net household income, adjusted for household composition using the modified OECD equivalence scale,^13^ which assigns a weight of 1 to the first adult, 0.5 to each additional adult, and 0.3 to each child. Equivalised income was then categorised into quintiles (1 = lowest, 5 = highest). IMD was measured separately across UK countries and categorized into quintiles (1 = most deprived, 5 = least deprived).^14^

### Analytic strategy

***Descriptive analyses*** were conducted to summarize sample characteristics and the distribution of adolescent mental health problems and health-risk behaviours. NEET prevalence across ages 16-24 years, the proportion ever experiencing NEET status, and NEET chronicity were described to characterize patterns of NEET status during late adolescence and early adulthood.

***Modified Poisson regression models*** with robust standard errors^15,16^ were used to estimate risk ratios (RRs) for the associations of adolescent mental health problems, health-risk behaviours, and co-occurring conditions with ever experiencing NEET status between ages 16 and 24 years.

***Proportional odds models*** were used to examine associations of adolescence exposure variables with NEET chronicity among respondents who had ever experienced NEET status. NEET chronicity was treated as an ordered categorical outcome, defined as one, two, and three or more NEET occasions. For each exposure of interest, the proportional odds assumption was formally tested by comparing a standard proportional odds model against a partial proportional odds model that allowed the exposure’s effect to vary across outcome thresholds, using a likelihood ratio test. When the assumption was violated (p<0.05), category-specific odds ratios were reported from the partial proportional odds model; otherwise, a common odds ratio across outcome thresholds was reported.

***Multilevel logistic regression models*** were built to examine the trajectory of NEET across ages 16-24 and to assess how adolescence exposures influence this trajectory. The age trajectory of NEET was first modelled without exposure variables. To account for potential nonlinearity in the age-NEET relationship, polynomial terms for age were included within the multilevel framework, with age centred at 16 years prior to analysis. Linear, quadratic, and cubic specifications were sequentially tested, and the optimal functional form was selected based on likelihood ratio tests (LRT), together with the Akaike Information Criterion (AIC) and Bayesian Information Criterion (BIC).

Subsequently, exposure-specific models were estimated separately to examine whether each exposure was associated with the initial level of NEET risk (intercept) and with the shape of the NEET trajectory over age (slope). To do so, an interaction term between the exposure and age was included in each model. Moderation effect was tested by comparing models with and without the interaction term using a LRT, with p<0.10 interpreted as evidence of moderation given the reduced power to detect interaction effects. Individual-level random intercepts were specified to account for the clustering of repeated observations within individuals across ages.

For all inferential analyses, each exposure was examined in a separate model to minimise overadjustment.^17^ All models were adjusted for sex, birth year, ethnicity, region of residence, household income, and area deprivation. Because exposures were modelled separately, missing values in one exposure did not affect the analytic sample for other exposures. Given this, together with the differing sample structures and modelling frameworks across the three

NEET outcomes, complete-case analysis was used throughout. To assess whether attrition in NEET observation could have biased estimates, negative binomial regression (adjusted for covariates) was used to examine whether adolescent exposures were associated with the number of waves in which NEET status was observed between ages 16 and 24 (Table S1). Data pre-processing and analyses were conducted in R version 4.5.2.

## RESULTS

### Sample characteristics

A total of 5,262 respondents were finally included in the analysis, of whom 49.3% were male and 23.8% were from minority ethnic backgrounds. Summary statistics for the study variables are presented in Table 1. Among the study sample, 1,474 participants (28.0%) experienced NEET status at least once between ages 16 and 24 years; most had a single NEET occasion (65.3%). The proportion classified as NEET increased with age and declined slightly after ages 21-22 years (Figure 1).

**Table 1.**
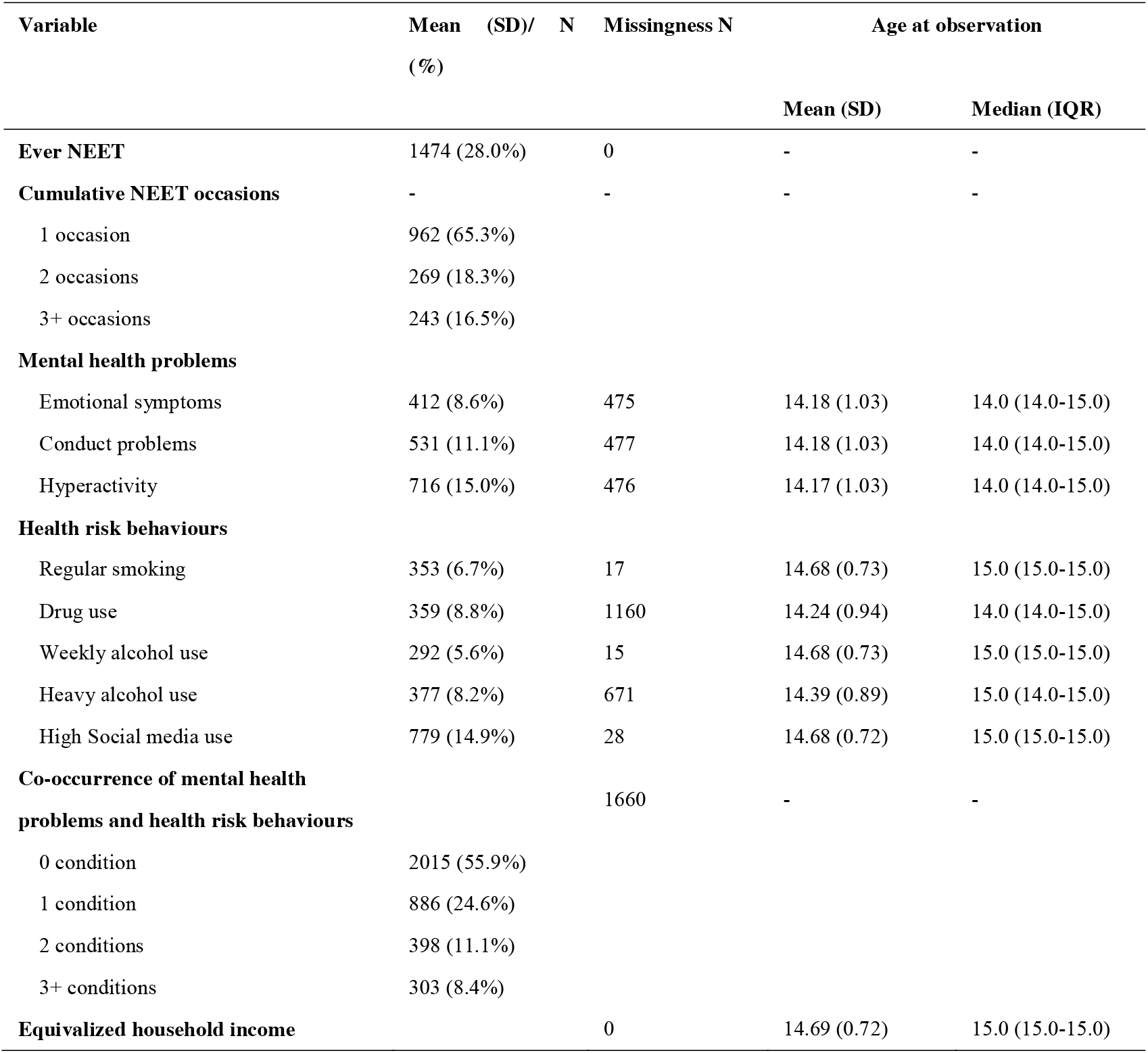
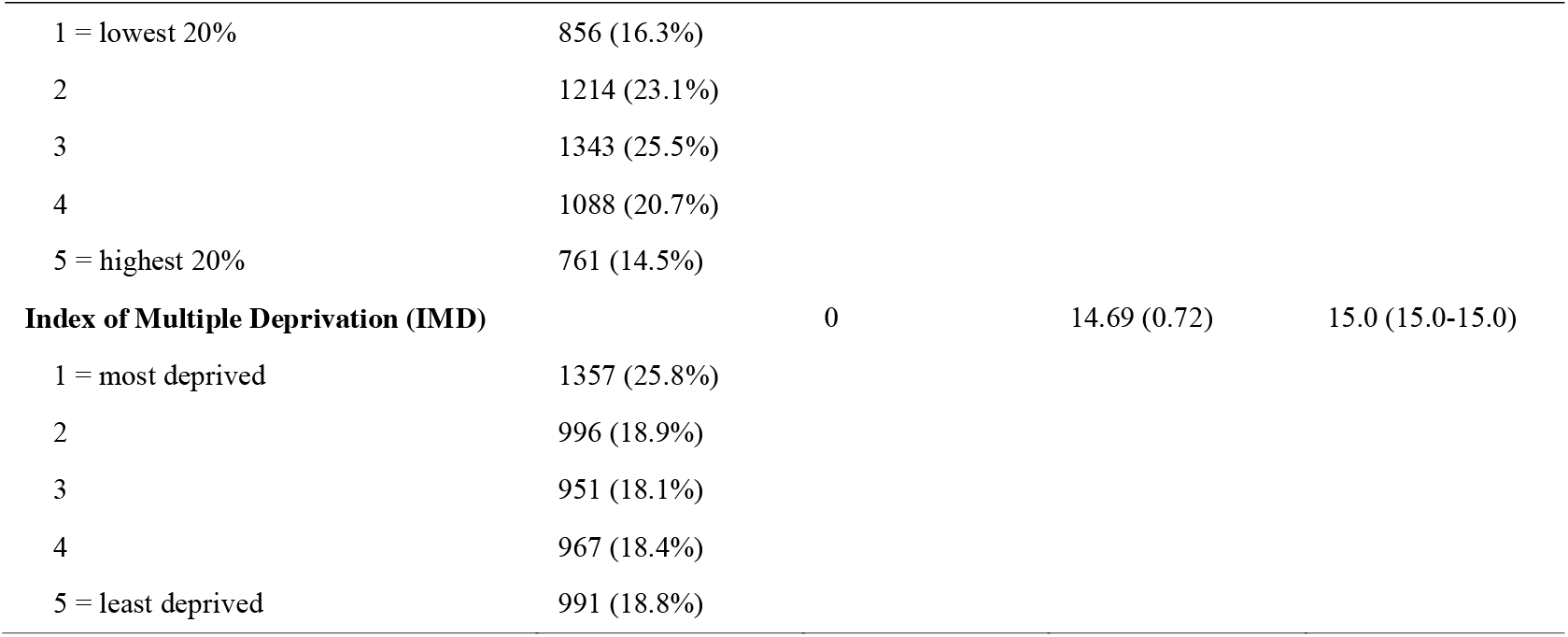
Descriptive characteristics of the analytic sample (N = 5,262)

**Figure 1.**
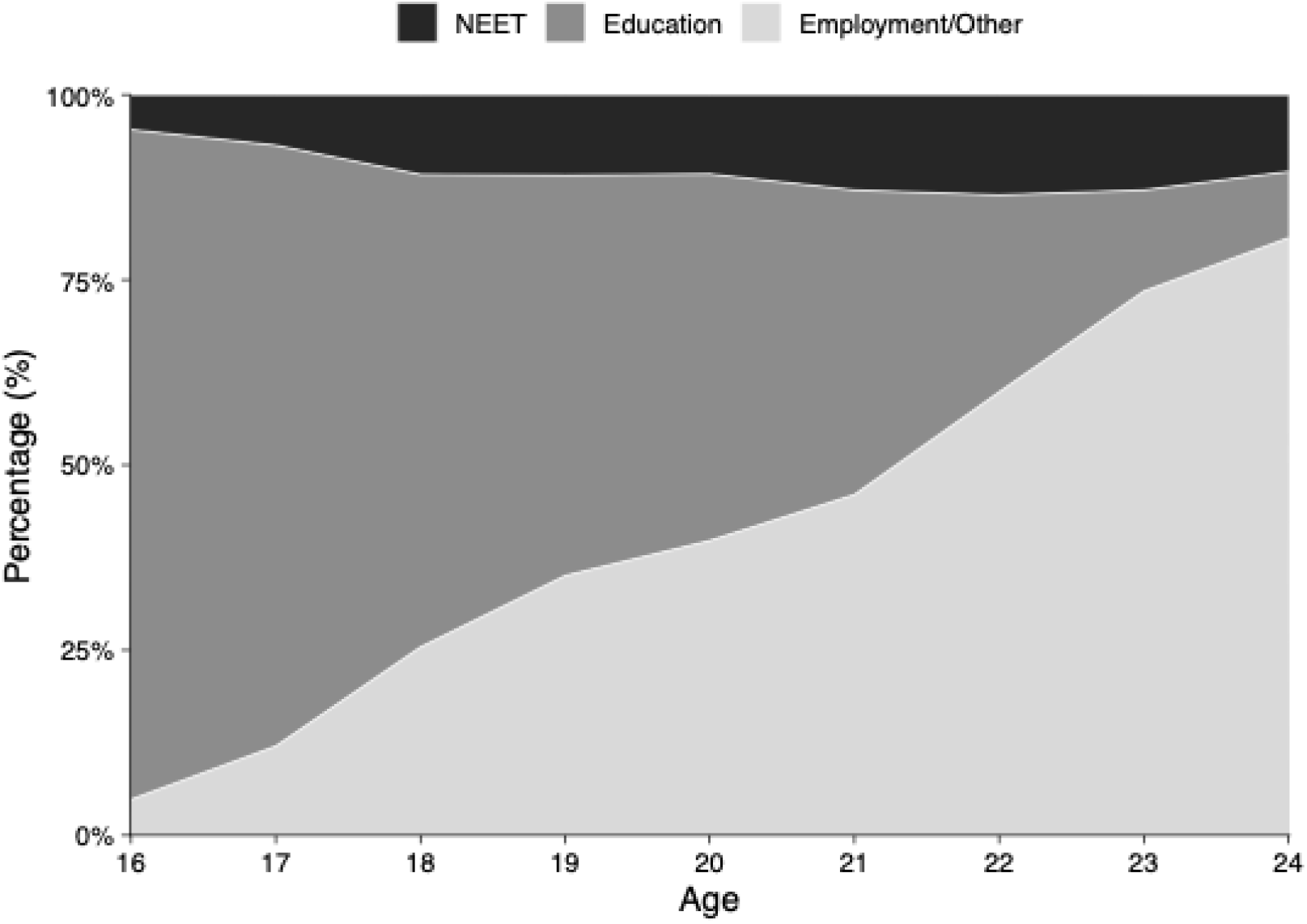
Distribution of NEET, education, and employment/other status across ages 16-24 years. ***Note*.** For descriptive purposes, non-NEET status was further classified into education and employment/other categories in this figure.

### Associations between adolescence exposures and ever-NEET status

Figure 2 and Table S2 present risk ratios (RRs) from modified Poisson regression models examining associations between adolescence exposures and ever experiencing NEET status.

**Figure 2.**
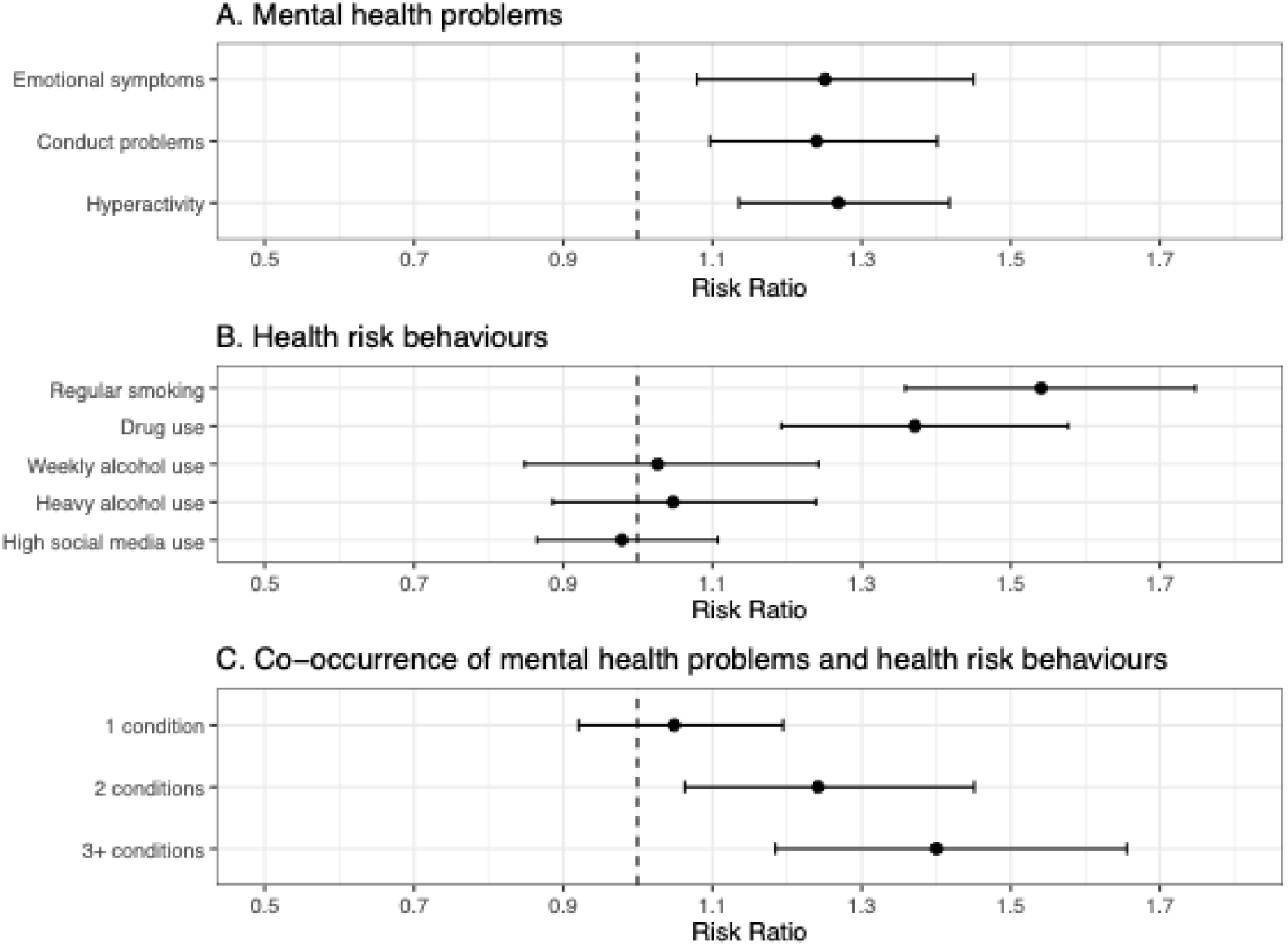
Associations between adolescence exposures and ever-NEET status across ages 16-24 years

Among mental health problems, all three domains were associated with increased risk of ever-NEET: hyperactivity (RR 1.27, 95% CI 1.14,1.42), emotional symptoms (RR 1.25, 95% CI 1.08,1.45), and conduct problems (RR 1.24, 95% CI 1.10,1.40), with broadly similar effect sizes.

Among health risk behaviours, regular smoking showed the strongest association (RR 1.54, 95% CI 1.36,1.75), followed by drug use (RR 1.37, 95% CI 1.19,1.58). Weekly alcohol use, heavy alcohol use, and high social media use were not associated with ever-NEET status.

Risk increased with the number of co-occurring mental health problems and health risk behaviours in a graded pattern: one condition (RR 1.05, 95% CI 0.92,1.20), two conditions (RR 1.24, 95% CI 1.06,1.45), and three or more conditions (RR 1.40, 95% CI 1.18,1.66).

### Associations between adolescence exposures and NEET chronicity

All exposures except the co-occurrence variable satisfied the proportional odds assumption, indicating that the association between each exposure and NEET chronicity was similar across different levels of NEET occasions. A single OR is therefore reported for each of these exposures (Figure 3, Table S3). Among mental health problems, emotional symptoms showed the strongest association with more NEET occasions (OR 1.98, 95% CI 1.39,2.83), followed by hyperactivity (OR 1.67, 95% CI 1.27,2.20) and conduct problems (OR 1.41, 95% CI 1.04,1.93). Among health risk behaviours, only regular smoking was associated with more NEET occasions (OR 1.64, 95% CI 1.19,2.28); no associations were observed for drug use, weekly and heavy alcohol use, and high social media use.

**Figure 3.**
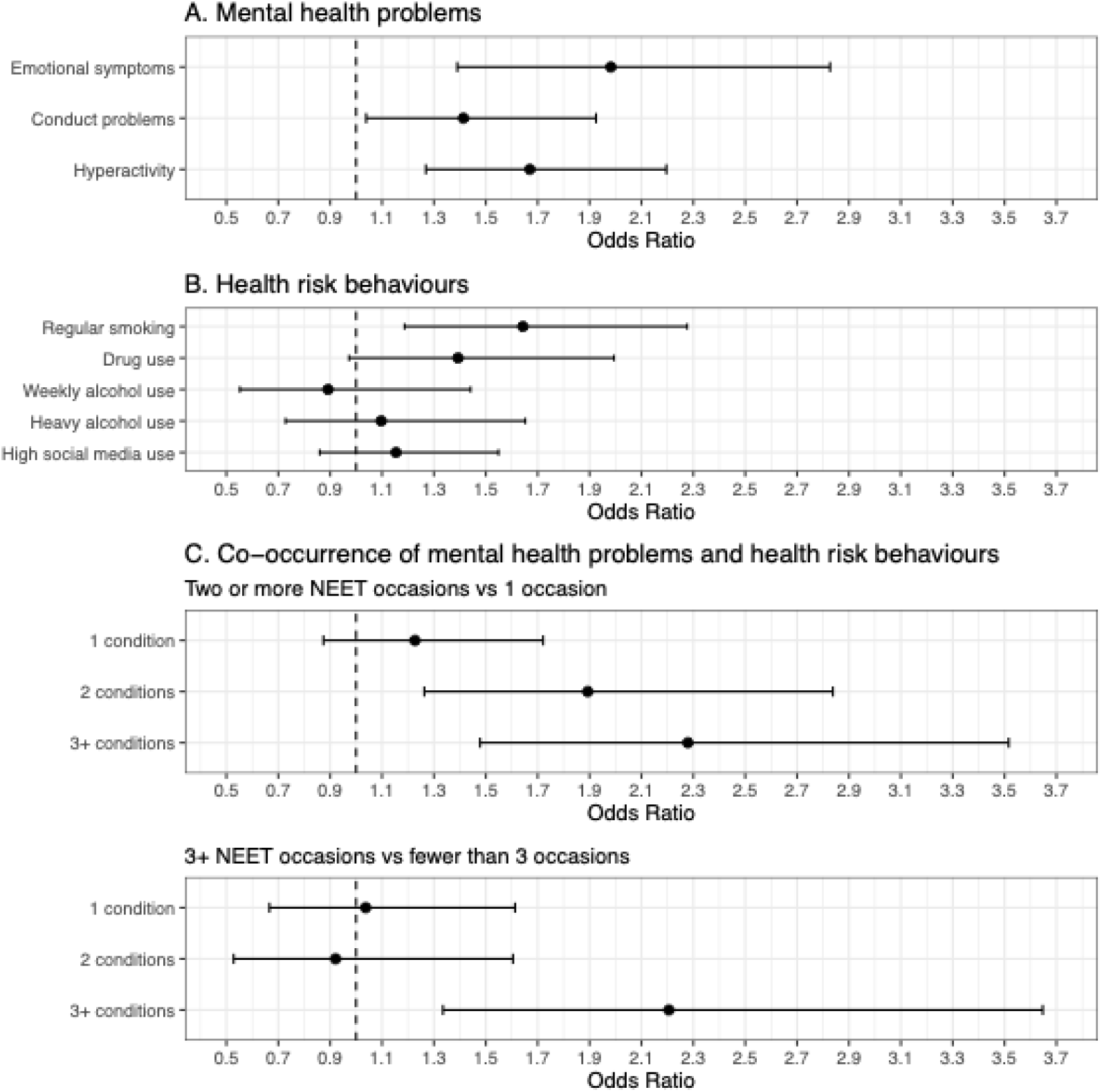
Associations between adolescence exposures and NEET chronicity across ages 16-24 years

The co-occurrence variable violated the proportional odds assumption (nominal test, p<0.05) and was modelled using a partial proportional odds specification, with separate ORs estimated at each threshold (Figure 3, Table S3). At the lower threshold (two or more vs one NEET occasion), odds increased with the number of co-occurring conditions: one condition (OR 1.23, 95% CI 0.87,1.72), two conditions (OR 1.89, 95% CI 1.26,2.84), three or more conditions (OR 2.28, 95% CI 1.48,3.52). At the higher threshold (three or more vs fewer than three occasions), only three or more co-occurring conditions were associated with greater NEET chronicity (OR 2.21, 95% CI 1.34,3.65).

### Age trajectory of NEET status across ages 16-24

The quadratic growth model demonstrated the best model fit for the data (see Table S4) and was therefore selected as the final model. The results of the exposure-specific mixed-effects models are summarized in Table S5 and illustrated in Figure 4. Consistent with the descriptive analysis, the predicted probability of NEET increased with age, declining slightly after 21-22 years (Figure S2). Distinct patterns of divergence in NEET trajectories between exposed and unexposed groups were observed across the different exposure variables.

**Figure 4.**
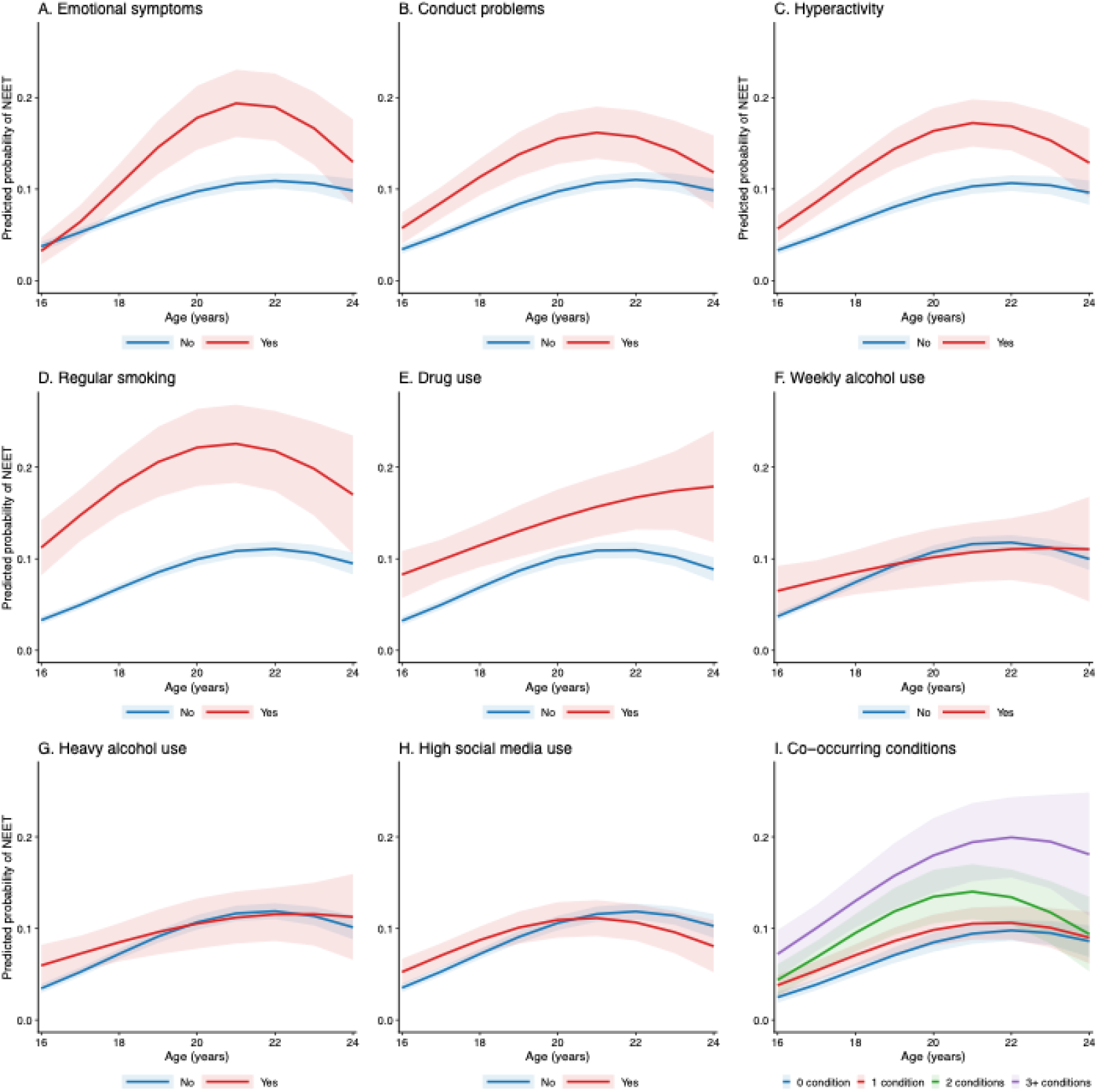
Predicted probability of NEET status across ages 16-24 for each adolescent mental health and health behaviour exposure, and their co-occurrence

For emotional symptoms (Figure 4A), the exposed and unexposed groups did not differ in NEET risk at baseline (age 16; OR 0.84, 95% CI 0.48,1.48, p=0.549); however, there was a moderation of the subsequent trajectory (LRT χ^2^=13.99, p<0.001): as age increased, the gap in NEET risk between the two groups widened, peaking at 21-22 years, before narrowing again.

For conduct problems and hyperactivity (Figure 4B, 4C), the exposed group showed higher NEET risk than the unexposed group at age 16 (conduct problems: OR 1.91, 95% CI 1.24,2.92, p=0.003; hyperactivity: OR 1.94, 95% CI 1.32,2.86, p=0.001), and this difference was largely maintained from ages 16 to 24, with no evidence of a moderation effect (conduct problems: χ^2^=2.48, p=0.290; hyperactivity: χ^2^=2.43, p=0.296).

For regular smoking (Figure 4D), the baseline difference between exposed and unexposed groups was the largest of all exposure variables (OR 4.88, 95% CI 3.20,7.43, p<0.001). The gap in NEET risk persisted across ages 16-24. There was evidence of moderation by age (χ^2^=5.48, p=0.065), with the gap widening slightly around ages 21-22.

For drug use (Figure 4E), the baseline difference was also substantial (OR 3.29, 95% CI 2.07,5.22, p<0.001), and the LRT indicated a difference in trajectory shape (χ^2^=6.16, p=0.046): while risk in the unexposed group declined after 21-22 years, risk in the exposed group increased monotonically with age.

For the two alcohol use variables (Figure 4F, 4G), exposed and unexposed groups differed at baseline (weekly alcohol use: OR 2.03, 95% CI 1.14,3.62, p=0.016; heavy alcohol use: OR 1.96, 95% CI 1.17,3.30, p=0.011). The LRTs suggested a potential moderation effect (weekly alcohol use: χ^2^=5.64, p=0.060; heavy alcohol use: χ^2^=5.75, p=0.056), with the gap in NEET risk between groups appearing to diminish with age.

For high social media use (Figure 4H), a baseline difference was also present (OR 1.64, 95% CI 1.11,2.42, p=0.013), and the LRT indicated moderation of the trajectory (χ^2^=11.31, p=0.004): after age 21, the rate of decline in NEET risk was faster in the exposed group than in the unexposed group.

For co-occurring conditions (Figure 4I), baseline NEET risk increased with the number of conditions (1 condition: OR 1.64, 95% CI 1.04,2.59, p=0.032; 2 conditions: OR 1.96, 95% CI 1.12,3.42, p=0.018; 3+ conditions: OR 3.70, 95% CI 2.12,6.47, p<0.001), but no moderation effect of the trajectory was found (LRT χ^2^=5.37, p=0.498).

## DISCUSSION

This study is the first to examine the associations of adolescence mental health problems, health risk behaviours, and their co-occurrence with three complementary NEET outcomes: ever experiencing NEET, NEET chronicity, and the age-related trajectory of NEET risk.

Although all exposures showed some evidence of association with NEET status, the strength and pattern of these associations varied across exposures and according to the outcome considered.

Among mental health problems, emotional, conduct and hyperactivity symptoms showed broadly similar associations with ever experiencing NEET, but differed in their associations with NEET chronicity and age-related trajectories. Emotional symptoms showed the strongest association with NEET chronicity, and their disadvantages emerged progressively over ages 16-24, whereas the disadvantage associated with conduct problems and hyperactivity was evident from age 16 and remained broadly stable. This difference might reflect the more immediate effects of externalising problems, such as conduct problems and hyperactivity, on school engagement through behavioural and disciplinary difficulties during mid-to-late adolescence, when most young people are still in education.^18^ By contrast, the consequences of emotional symptoms may become more apparent as young people navigate increasingly demanding educational and employment transitions as they move into early adulthood.

Among health-risk behaviours, regular smoking emerged as the most consistently relevant exposure. Drug use showed a similarly strong association with ever experiencing NEET, but not with chronicity, which may have been influenced by the relatively small sample size. However, unlike non-users, whose NEET risk declined after ages 21-22, risk among drug users continued to rise with age, suggesting a greater likelihood of prolonged NEET involvement during the transition into the labour market, consistent with previous evidence linking early substance use disorders to longer periods of NEET involvement in adulthood.^19^ The two alcohol use exposures appeared to have only a limited relationship with NEET: neither was associated with ever experiencing NEET or with chronicity, and although a disadvantage in NEET risk was evident at age 16, it tended to diminish over time. High social media use showed a similarly limited relationship with NEET. Taken together, smoking and drug use showed the most robust and persistent associations with NEET risk, whereas alcohol use appeared to be comparatively weak and unstable correlate, consistent with previous evidence.^5^ Social media use, however, is a multidimensional construct, and frequency or duration alone is not consistently relate to adverse outcomes,^20^ with associations potentially varying by the nature and context of use.

In line with previous evidence,^21,22^ co-occurring mental health problems and health-risk behaviours showed a graded association with NEET risk across several outcomes: risk appeared to rise with the number of co-occurring conditions, both for ever experiencing NEET and for the age-16 baseline risk in the trajectory model, where this graded disadvantage remained stable across ages 16-24 with no evidence of moderation by age. A similar graded pattern was also evident for NEET chronicity at the lower threshold (distinguishing one from two or more NEET occasions). However, at the higher threshold, only three or more co-occurring conditions were associated with the risk of having three or more NEET occasions, suggesting that a threshold rather than a graded pattern may better characterize the association with the most severe and recurrent NEET experiences. These particularly high-risk young people may benefit more from early, coordinated support that addresses multiple difficulties together, rather than interventions targeting a single problem in isolation.

This study has several limitations. First, to maximise the use of available information, exposure variables were measured using the latest available observation in the youth survey. This approach could have introduced heterogeneity in the timing of exposure measurement relative to the onset of NEET risk, but the age at which exposures were assigned was highly concentrated across all exposures (median 14-15 years, IQR 14-15), suggesting that any resulting heterogeneity was likely limited. Second, as with most longitudinal studies, attrition was unlikely to be random: respondents with conduct problems, hyperactivity, regular smoking, or alcohol use had fewer observations of NEET status across ages 16-24, potentially underestimating the associations of these exposures with NEET risk. Third, the measure of co-occurrence captured only the number of co-occurring conditions, rather than the specific combinations of mental health problems and health-risk behaviours present. Different combinations of conditions might carry different levels of risk, but unpacking this multidimensionality would need greater sample sizes. Future studies with larger samples could apply approaches such as multilevel analysis of individual heterogeneity and discriminatory accuracy (MAIHDA) to examine whether specific combinations of conditions are associated with differential risk. Fourth, this study examined a specific range of adolescent mental health problems and health-risk behaviours that were available in this large survey. Future studies could examine other potentially relevant exposures, such as problematic social media use, as well as finer-grained health and diagnostic measures if available. Finally, our measures of NEET chronicity did not capture the timing or sequencing of NEET occasions, due to limited power to examine diverse potential NEET typologies and transitions. Future research using larger scale data such as administrative records could examine the timing, continuity, and typologies of NEET experiences in greater detail.

## CONCLUSIONS

Mental health problems and health-risk behaviours in adolescence are associated with the risk of being NEET between ages 16 and 24. Emotional, conduct, and hyperactivity problems differed in the timing and persistence of their link to NEET, suggesting that domains whose associated risk emerges later (e.g., emotional symptoms) should not be deprioritised for early intervention. Smoking and drug use showed more consistent relationships with NEET than alcohol use and social media use, suggesting they may be more useful early markers of NEET risk. NEET risk increased with the number of co-occurring conditions, and elevated risk for the most severe and recurrent NEET experiences (three or more occasions) was observed only with three or more co-occurring conditions, underscoring the particular vulnerability of adolescents with multiple concurrent risk indicators.

## Data Availability

The data for this study are available from the UK Data Service.

## Availability of data and materials

The data for this study are available from the UK Data Service.

## Ethics approval and consent to participate

Ethical approval for these analyses were received from the UCL Institute of Education research ethics committee (REC 2428). Study members provided active consent to be included in the study.

## Authors’ contributions

ZL - Conceptualisation, Methodology, Formal Analysis, Data Curation, Writing - Original Draft, Visualisation;

JW, NC - Conceptualisation, Writing - Review & Editing, Supervision, Funding;

PP- Conceptualisation, Methodology, Writing - Review & Editing, Supervision, Funding. All authors read and approved the final manuscript.

## Acknowledgments

Thank you to the participants of the UKHLS, and to the teams at University of Essex for curating the study and making the data available for research.

