## Supplementary Material for "Mental health and health behaviours in adolescence and risk of being NEET from ages 16 to 24: longitudinal findings from the UKHLS"

**Figure S1**

*Flow diagram showing analytic sample selection*

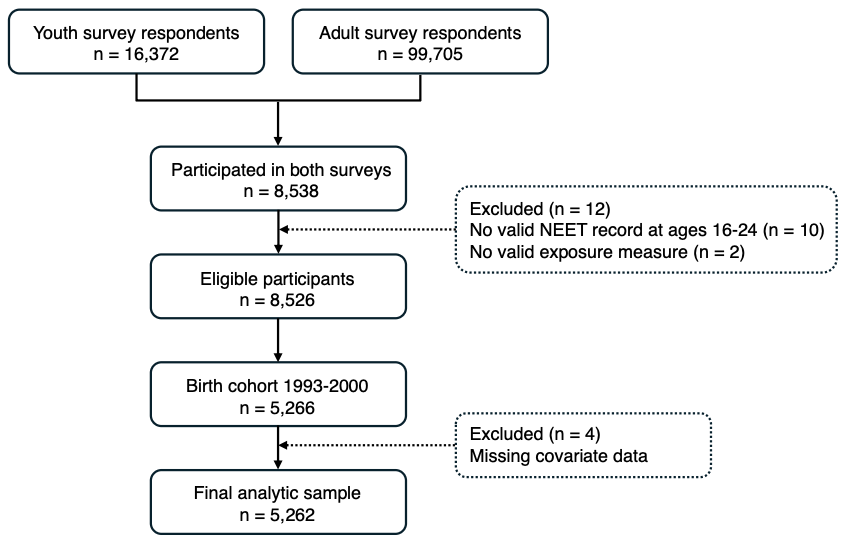

**Table S1**

*Number of waves NEET status was observed during ages 16-24, and its association with adolescence exposures*

| ***Panel A: Distribution of number of waves observed*** | | |
| --- | --- | --- |
| **Number of waves observed** | **N** | **%** |
| 1 | 697 | 13.2 |
| 2 | 597 | 11.3 |
| 3 | 603 | 11.5 |
| 4 | 563 | 10.7 |
| 5 | 554 | 10.5 |
| 6 | 554 | 10.5 |
| 7 | 585 | 11.1 |
| 8 | 583 | 11.1 |
| 9 | 526 | 10.0 |
| **Total** | **5,262** | **100.0** |
| ***Panel B: Association between adolescence exposures and number of waves observed*** | | |
| **Exposure** | **IRR (95%CI)** | ***p* value** |
| Emotional symptoms | 0.99 (0.94, 1.05) | 0.797 |
| Conduct problems | 0.94 (0.90, 0.99) | 0.027 |
| Hyperactivity | 0.91 (0.87, 0.95) | <0.001 |
| Regular smoking | 0.87 (0.82, 0.92) | <0.001 |
| Drug use | 0.95 (0.90, 1.01) | 0.114 |
| Weekly alcohol use | 0.92 (0.86, 0.99) | 0.019 |
| Heavy alcohol use | 0.94 (0.89, 1.00) | 0.034 |

***Note.*** An overdispersion test (dispersion = 1.41, p<0.001) indicated that negative binomial regression was preferred over Poisson regression. Results are reported as incidence rate ratios (IRRs) with 95% confidence intervals; an IRR below 1 indicates that a given exposure was associated with fewer observed waves.

**Table S2**

*Regression results for adolescence exposures and ever-NEET status*

| **Exposure** | **N_ind_** | **RR (95%CI)** |
| --- | --- | --- |
| **Mental health problems** |  |  |
| Emotional symptoms | 4787 | **1.25 (1.08,1.45)** |
| Conduct problems | 4785 | **1.24 (1.10,1.40)** |
| Hyperactivity | 4786 | **1.27 (1.14,1.42)** |
| **Health risk behaviours** |  |  |
| Regular smoking | 5245 | **1.54 (1.36,1.75)** |
| Drug use | 4102 | **1.37 (1.19,1.58)** |
| Weekly alcohol use | 5247 | 1.03 (0.85,1.24) |
| Heavy alcohol use | 4591 | 1.05 (0.89,1.24) |
| High Social media use | 5234 | 0.98 (0.87,1.11) |
| **Co-occurrence of mental health problems and health risk behaviours** | 3602 |  |
| 1 condition |  | 1.05 (0.92,1.20) |
| 2 conditions |  | **1.24 (1.06,1.45)** |
| 3+ conditions |  | **1.40 (1.18,1.66)** |

***Note.*** RR = Risk ratio.

**Table S3**

*Regression results for adolescence exposures and NEET chronicity (number of NEET occasions)*

| **Exposure** | **N_ind_** | **OR (95%CI)** |
| --- | --- | --- |
| **Mental health problems** |  |  |
| Emotional symptoms | 1323 | **1.98 (1.39,2.83)** |
| Conduct problems | 1323 | **1.41 (1.04,1.93)** |
| Hyperactivity | 1323 | **1.67 (1.27,2.20)** |
| **Health risk behaviours** |  |  |
| Regular smoking | 1467 | **1.64 (1.19,2.28)** |
| Drug use | 1156 | 1.39 (0.97,1.99) |
| Weekly alcohol use | 1469 | 0.89 (0.55,1.44) |
| Heavy alcohol use | 1296 | 1.10 (0.73,1.65) |
| High Social media use | 1459 | 1.15 (0.86,1.55) |
| **Co-occurrence of mental health problems and health risk behaviours** | 995 |  |
| Two or more NEET occasions vs 1 occasion |  |  |
| 1 condition |  | 1.23 (0.87,1.72) |
| 2 conditions |  | **1.89 (1.26,2.84)** |
| 3+ conditions |  | **2.28 (1.48,3.52)** |
| 3+ NEET occasions vs fewer than 3 occasions |  |  |
| 1 condition |  | 1.04 (0.67,1.61) |
| 2 conditions |  | 0.92 (0.53,1.61) |
| 3+ conditions |  | **2.21 (1.34,3.65)** |

***Note.*** OR = odds ratio.

**Table S4**

*Comparison of the linear, quadratic, and cubic growth models*

| **Model** | **AIC** | **BIC** | **LRT χ^2^** | **LRT *p*** |
| --- | --- | --- | --- | --- |
| Linear growth | 14226 | 14323 | - | - |
| Quadratic growth | 14128 | 14234 | 99.247 | <0.001 |
| Cubic growth | 14129 | 14243 | 1.868 | 0.172 |

***Note.*** AIC = Akaike Information Criterion; BIC = Bayesian Information Criterion; LRT = likelihood ratio test. LRT χ^2^ and p-values are from chi-square difference tests comparing each model to the preceding specification. The quadratic model provided the best fit, with a significant improvement over the linear model (LRT χ^2^ = 99.25, p < 0.001), while the cubic term did not yield further improvement (LRT χ^2^ = 1.87, p = 0.172). The quadratic specification was therefore selected for all subsequent analyses.

**Table S5**

*Moderation of age-related NEET trajectories by mental health and health behaviours during adolescence*

| **Exposure** | **N_obs_** | **N_ind_** | **LRT χ^2^** | **LRT *p*** | **Term** | **OR (95%CI)** | ***p* value** |
| --- | --- | --- | --- | --- | --- | --- | --- |
| Emotional symptoms | 23370 | 4787 | 13.99 | **<0.001** | Emotional symptoms | 0.84 (0.48,1.48) | 0.549 |
|  |  |  |  |  | **Age interaction (linear)** | **1.61 (1.21,2.14)** | **0.001** |
|  |  |  |  |  | **Age interaction (quadratic)** | **0.95 (0.92,0.98)** | **0.004** |
| Conduct problems | 23363 | 4785 | 2.48 | 0.290 | **Conduct problems** | **1.91 (1.24,2.92)** | **0.003** |
|  |  |  |  |  | Age interaction (linear) | 1.08 (0.86,1.35) | 0.535 |
|  |  |  |  |  | Age interaction (quadratic) | 0.99 (0.96,1.01) | 0.311 |
| Hyperactivity | 23369 | 4786 | 2.43 | 0.296 | **Hyperactivity** | **1.94 (1.32,2.86)** | **0.001** |
|  |  |  |  |  | Age interaction (linear) | 1.12 (0.91,1.38) | 0.283 |
|  |  |  |  |  | Age interaction (quadratic) | 0.98 (0.96,1.01) | 0.179 |
| Regular smoking | 25480 | 5245 | 5.48 | **0.065** | **Regular smoking** | **4.88 (3.20,7.43)** | **<0.001** |
|  |  |  |  |  | Age interaction (linear) | 0.92 (0.72,1.17) | 0.492 |
|  |  |  |  |  | Age interaction (quadratic) | 1.00 (0.97,1.03) | 0.970 |
| Drug use | 20358 | 4102 | 6.16 | **0.046** | **Drug use** | **3.29 (2.07,5.22)** | **0.000** |
|  |  |  |  |  | **Age interaction (linear)** | **0.73 (0.57,0.94)** | **0.015** |
|  |  |  |  |  | **Age interaction (quadratic)** | **1.04 (1.01,1.07)** | **0.023** |
| Weekly alcohol use | 25481 | 5247 | 5.64 | **0.060** | **Weekly alcohol use** | **2.03 (1.14,3.62)** | **0.016** |
|  |  |  |  |  | Age interaction (linear) | 0.72 (0.52,1.00) | 0.053 |
|  |  |  |  |  | Age interaction (quadratic) | 1.03 (0.99,1.08) | 0.137 |
| Heavy alcohol use | 22517 | 4591 | 5.75 | **0.056** | **Heavy alcohol use** | **1.96 (1.17,3.30)** | **0.011** |
|  |  |  |  |  | Age interaction (linear) | 0.75 (0.57,1.00) | 0.053 |
|  |  |  |  |  | Age interaction (quadratic) | 1.03 (0.99,1.06) | 0.136 |
| High Social media use | 25426 | 5234 | 11.31 | **0.004** | **High Social media use** | **1.64 (1.11,2.42)** | **0.013** |
|  |  |  |  |  | Age interaction (linear) | 0.89 (0.72,1.10) | 0.271 |
|  |  |  |  |  | Age interaction (quadratic) | 1.00 (0.97,1.03) | 0.889 |
| Co-occurrence conditions | 18028 | 3602 | 5.37 | 0.498 | **1 condition** | **1.64 (1.04,2.59)** | **0.032** |
|  |  |  |  |  | **2 conditions** | **1.96 (1.12,3.42)** | **0.018** |
|  |  |  |  |  | **3+ conditions** | **3.70 (2.12,6.47)** | **<0.001** |
|  |  |  |  |  | Age interaction (1 condition, linear) | 0.91 (0.72,1.16) | 0.457 |
|  |  |  |  |  | Age interaction (1 condition, quadratic) | 1.00 (0.98,1.03) | 0.762 |
|  |  |  |  |  | Age interaction (2 conditions, linear) | 1.07 (0.79,1.43) | 0.668 |
|  |  |  |  |  | Age interaction (2 conditions, quadratic) | 0.98 (0.95,1.02) | 0.374 |
|  |  |  |  |  | Age interaction (3+ conditions, linear) | 0.93 (0.69,1.26) | 0.639 |
|  |  |  |  |  | Age interaction (3+ conditions, quadratic) | 1.01 (0.97,1.04) | 0.762 |

***Note.*** N_obs_ = number of observations; N_ind_ = number of respondents; LRT χ^2^ = likelihood ratio test statistic, comparing the main-effects model to the interaction model; LRT *p* = likelihood ratio test p-value; OR = odds ratio from the interaction model; CI = confidence interval. All models are mixed-effects logistic regressions with random intercepts for individuals adjusted for sex, birth year, ethnicity, region of residence, household income, and area deprivation.

**Figure S2**

*Predicted probability of NEET from age 16 to 24*

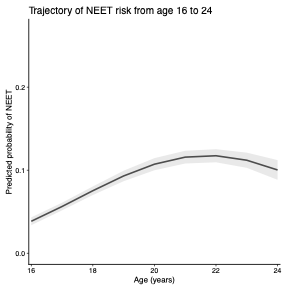
